# *Thomasclavelia ramosa* is a Signature of Gut Dysbiosis associated with Alcohol-Related Hepatocellular Carcinoma: A First Microbial Culturomics Study

**DOI:** 10.1101/2024.08.19.24312231

**Authors:** Reham Magdy Wasfy, Anissa Abdoulaye, Patrick Borentain, Babacar Mbaye, Maryam Tidjani Alou, Aurelia Caputo, Claudia Andrieu, Giovanna Mottola, Anthony Levasseur, Matthieu Million, Rene Gerolami

**Affiliations:** IHU Méditerranée Infection, Marseille, France; MEPHI, Aix-Marseille Université, Marseille, France; Unité hépatologie, Hôpital de la Timone, Marseille, France; Assistance Publique-Hôpitaux de Marseille (APHM), Marseille, France; Laboratoire de Biochimie, Hôpital de la Timone, APHM, 13005 Marseille, France; C2VN, INSERM 1263, INRAE 1260, Team 5, Aix-Marseille Université, 13005 Marseille, France

**Keywords:** *Thomasclavelia ramosa*, *Enterocloster bolteae*, *Mediterraneibacter gnavus*, Alcoholic-related liver disease, Hepatocellular carcinoma, Gut microbiome, Microbial culturomics, 16S rRNA sequencing, Cancer

## Abstract

**Background:** Gut microbiota alteration is implicated in the pathogenesis of alcoholic liver disease (ALD) and HCC. No study has characterized the dysbiosis associated with ALD by microbial culturomics, an approach that certifies viability and allows the characterization of pathobiont strain candidates.

**Methods:** A case-control study was conducted on patients with ALD without HCC (ALD-NoHCC) (n=16), ALD with HCC (ALD-HCC) (n=19), and controls (n=24). 16S rRNA amplicon sequencing and microbial culturomics were used as complementary methods for gut microbiome profiling.

**Results:** By microbial culturomics, *Thomasclavelia ramosa* was the most enriched and detected in all ALD samples (100%), while it was cultivated in only a small proportion of controls (20%, p < 0.001). By 16S rRNA amplicon sequencing and 3-groups linear discriminant analysis, *T. ramosa* was increased explicitly in the ALD-HCC group (LDA-score > 5, p < 0.05).

**Conclusions:** *T. ramosa,* identified by culturomics and 16 rRNA sequencing, is associated with ALD and ALD-HCC. Alongside the recently reported in vitro genotoxicity of this species in colorectal cancer, this species has been identified as a candidate oncobiont in ALD-HCC.

**Highlights:**

- The gut microbiota signature of ALD and ALD-HCC was explored by microbial culturomics and 16S amplicon sequencing
- By culturomics, *T. ramosa* was the most enriched and cultured from all included ALD patients, but in only 20% of controls (p < 0.05).
- *T. ramosa* was significantly associated with alcohol-related HCC by 16S sequencing.
- *T. ramosa* is identified as a putative oncobiont associated with ALD-HCC, thus opening new avenues for diagnosis and treatment.

**Graphical abstract:** 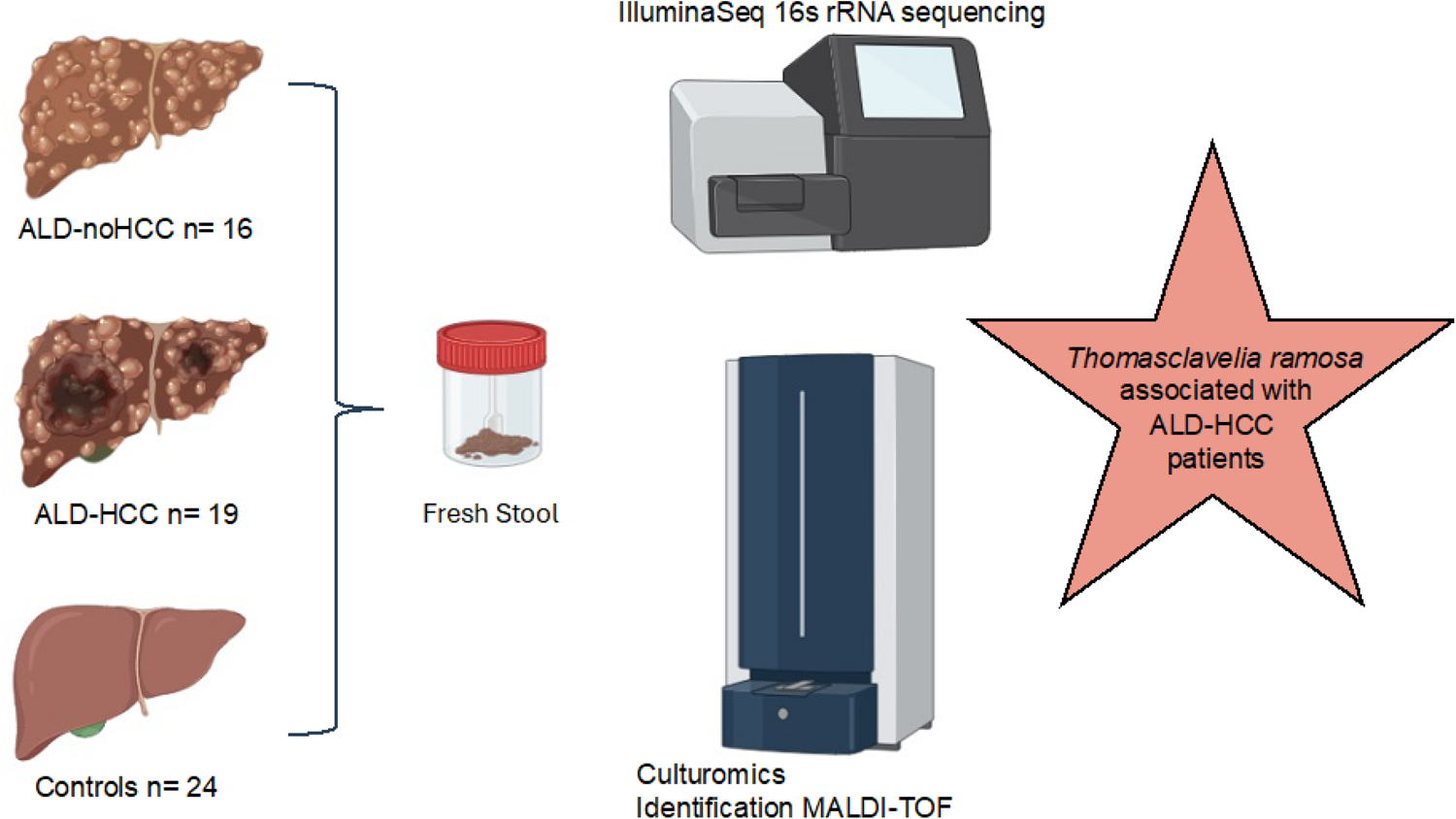

## Introduction

Hepatocellular carcinoma (HCC) is the most common form of liver cancer (90% of cases), primarily associated with chronic liver disease. Its major risk factors include chronic viral hepatitis infection (HBV, HCV), Alcoholic Liver Disease (ALD), and Metabolic dysfunction-associated Steato-Hepatitis (MASH) (1). Concerning the incidence and mortality in the world, liver cancer ranks sixth and third place across diverse cancers, which has caused a great cancer burden globally (2).

The link between alcohol, liver disease, and cancer is well-established. Its mechanism would include direct toxicity of alcohol on the liver, but persistent instrumental factors are suspected, as the evolution of the disease is not reversible upon withdrawal (3). Among persistent instrumental factors, the gut liver axis and gut microbiota are potential candidates to explain the persistence of a vicious circle that would explain the persistent excess risk up to 10 years after weaning (4). Recent studies suggest that alcohol dependence syndrome and alcoholic liver disease are both associated with gut microbiota alteration with distinct features (5, 6). This indicates that a specific gut dysbiosis may enhance cirrhosis and hepatocarcinogenesis by the gut-liver axis. Indeed, some data from human studies (7–12) and animal experimental models (13–18) indicate that HCC occurrence is related to gut microbiota, and treatment with broad-spectrum antibiotics decreases HCC tumor growth in mice (19).

Several metagenomic studies have investigated gut dysbiosis in patients with ALD based on 16S rRNA amplicon sequencing (20–22) or shotgun (whole-genome) sequencing (6). However, no study described gut dysbiosis in patients with ALD or HCC based on the culturomics approach. We found a study reporting cultured microbial counts but with minimal microbial taxonomic accuracy and evidencing an *Escherichia coli* enrichment (7).

Microbial culturomics is a new - omics strategy developed in our center as a high-throughput culture method based on Matrix-Assisted Laser Desorption Ionization Time-Of-Flight Mass Spectrometry (MALDI-TOF MS) and diversified physicochemical culture conditions mimicking the natural microenvironment (23), able to identify several new species (24) and complementary to metagenomics (25). MALDI-TOF MS can identify hundreds of colonies per hour with unprecedented microbial taxonomic accuracy (26). Several teams have successfully reproduced this approach to explore the human microbiota composition (27), notably by illuminating the metagenomic microbial dark matter (28). Culturomics has the unparalleled advantage of ascertaining viable microbial strains. In contrast, metagenomics and sequencing approaches may have the ‘running after ghosts’ bias, i.e., identify only DNA sequences unrelated to viable bacteria (29).

Furthermore, our findings demonstrated that this microbial culturomics complements metagenomics when assessing a dysbiotic signature in a clinical context (severe acute malnutrition (30), necrotizing enterocolitis (31), and HIV (32)). Our team applied fungal and bacterial culturomics in the context of liver diseases, demonstrating the importance of ethanol-producing yeast and bacteria in MASH and HBV-associated liver disease (33, 34). Some results were expected in the most recent metagenomic studies, such as the association of *Limosilactobaccilus fermentum* with MASH (35). In contrast, only fungal and bacterial culturomics identified certain yeast species of the genus *Candida* and *Pichia* and bacterial species of the genus *Enterocloster* associated with MASH (33, 34). *Enterocloster* was also associated with liver disease in the context of HBV infection (36). Again, it was found only by culturomics. Different biases may explain the discrepancies between DNA sequencing and culturomics, making these two approaches complementary (25). However, to our knowledge, no study has yet characterized microbiota associated with alcoholic liver disease using microbial culturomics.

Accordingly, this study aims to characterize the microbial signature in patients with ALD and ALD-associated HCC using both culturomics and large-scale sequencing (v3v4 region 16S rRNA amplicon sequencing).

## Methods

### Study Design and ethics statement

A case-control study was carried out in the Hepatology Department of Marseille University Hospital (south-eastern France), Marseille, France, according to STROBE statement guidelines (37) from January to June 2022. The HEPATGUT study was approved by the local ethics committee of the Institut Hospitalo-Universitaire Méditerranée Infection, Marseille, France (IHUMI, 2020-004), approved by the Protection of Persons Committee (Approval No. CPP: 21.04391.000046—21075), and carried out according to the 2013 Declaration of Helsinki (World Medical Association, 2013) (38). Patient consent (non-opposition) was obtained according to French regulations.

### Study Population

The HEPATGUT study is a case-control study focusing on gut microbiota alteration in hepatic diseases. Previously published results focused on HBV (36) and MASH (33, 34). The present study included 35 patients with ALD and 24 controls (CTL). All 59 provided a fecal sample analyzed using 16S rRNA sequencing, and 21 were analyzed by culturomics (11 ALD and 10 CTL). This is explained by the fact that the workload is much higher for culturomics (6 weeks per sample) than for 16S rRNA sequencing. Healthy controls without chronic diseases or on regular medications were recruited. Age, gender, weight, height, and alcohol consumption were collected. Patients with ALD were diagnosed according to the European Association for the Study of the Liver (EASL) guidelines (39).

The main exclusion criteria were probiotics/antibiotics intake in the previous month before the collection of stools, participants under 18 years old, and pregnancy. To clarify gut microbiota signature according to HCC, we divided the ALD group into ALD without HCC (ALD-NoHCC) and ALD with HCC (ALD-HCC). The diagnosis of HCC was made according to the EASL guidelines (40). Controls (CTL) were recruited as previously reported (34, 36). The groups were matched with age and body mass index (BMI).

### Clinical measurements

Liver stiffness measurements in cirrhotic patients were conducted using a FibroScan® instrument (Echosens, Paris, France). Measurements with more than ten successful acquisitions were obtained (with a rate of > 60% and an interquartile range of < 30%). In addition, routine biochemistry, including prothrombin index (PT), platelets count (PLT), total bilirubin (TBIL), serum albumin (ALB), alanine aminotransferase (ALT), aspartate aminotransferase (AST), gamma-glutamyl transferase (GGT), alkaline phosphatase (ALP) and serum creatinine, were measured. The fecal samples from all participants were collected and stored at −80° C until used, as described in our previous study (36).

### Microbial culturomics

Twenty-one stool samples (11 samples from patients with ALD and ten samples from controls) were cultured according to the culturomics approach previously established in our laboratory (25)This number of samples was chosen because microbial culturomics is a time-consuming approach. A full-time person takes approximately six weeks to analyze each sample. In addition, the culturomics study is intended to be an exploratory study to gain insights into cultured microbiota in patients with ALD and HCC. Fast culturomics was applied using four culture conditions, as comprehensively detailed by Naud et al. (41). We applied the same culturomics methodology described in our previous study (36). A subculture of the isolated colonies was performed for purification. The colonies were then identified using MALDI-TOF MS (Bruker Daltonics, Bremen, Germany) according to the manufacturer’s instruction (26).

### 16S rDNA gene sequencing

PCR amplification of the bacterial 16S rRNA gene V3-V4 region was performed using primers (FwOvAd_341F:5’TCGTCGGCAGCGTCAGATGTGTATAAGAGACAGCCTACGGGNGG CWGCAG3’; RevOvAd_785R:5’GTCTCGTGGGCTCGGAGATGTGTATAAGAGACAGGACTACHVGGGTATCTAATCC3’) with a Illumina MiSeq engine as being described in details in our previous study (36).

### Bioinformatics analysis

The raw sequencing data for all samples were deposited into the NCBI Sequence Read Archive database (https://www.ncbi.nlm.nih.gov/bioproject/?term=PRJEB62828). Noisy sequencing data were excluded, and chimeric sequences were identified and removed by Chimera Slayer. The clean data were clustered into operational taxonomic units (OTUs) at the 97% similarity threshold using the UCLUST algorithm after the removal of singletons. The alpha and beta diversity were calculated using the MicrobiomeAnalyst database (https://www.microbiomeanalyst.ca/; Accessed on 30 September 2023) based on the profile of the relative abundance of OTUs in a single sample. Alpha diversity was evaluated through three indexes (Chao1, Simpson, and Shannon) to compare bacterial richness and diversity across samples. Principal Coordinates analysis (PCoA) was conducted using Bray-Curtis dissimilarity to visualize the microbial structure and distribution among the samples.

Permutational multivariate analysis of variance (PERMANOVA) was used to assess beta diversity. Linear discriminant analysis (LDA) effect size (LEfSe) used the Kruskal-Wallis rank sum test combined with LDA to detect features with significantly different abundances of various taxa among groups was done with the Microbiome Analyst Platform (https://www.microbiomeanalyst.ca/; Accessed 30 September 2023). A log LDA score >2 was the threshold for discriminating between groups.

### Statistical analysis

Normal distribution was determined using either D’Agostino-Pearson or the Kolmogorov-Smirnov test. One-way analysis of variance was performed to compare continuous variables between two groups. Differences between groups were determined by using the non-parametric Kruskal-Wallis and Mann-Whitney U test. Spearman rank correlation was used to calculate the relationship between microbial makers and clinical indicators. The two-sided Fisher’s exact test was used for significant detection frequency difference. All statistical analyses were calculated using GraphPad Prism Software for Windows (GraphPad Software, San Diego, CA, USA) (version 9.0). P values less than 0.05 were considered significant differences.

## RESULTS

### Characteristics of the study participants

After a strict inclusion and exclusion process, 59 participants were enrolled in our study, including 35 patients with ALD and 24 controls. The ALD group was subdivided into ALD without HCC (ALD-NoHCC, n=16) and with HCC (ALD-HCC, n=19) (Figure 1). The clinical characteristics of the groups are shown in Table 1. The three groups were relatively well-balanced in sample size, allowing relevant comparisons. Compared to controls, ALD was associated with male gender, high blood pressure, and smoking, as expected (Table 1).

**Figure. 1.**
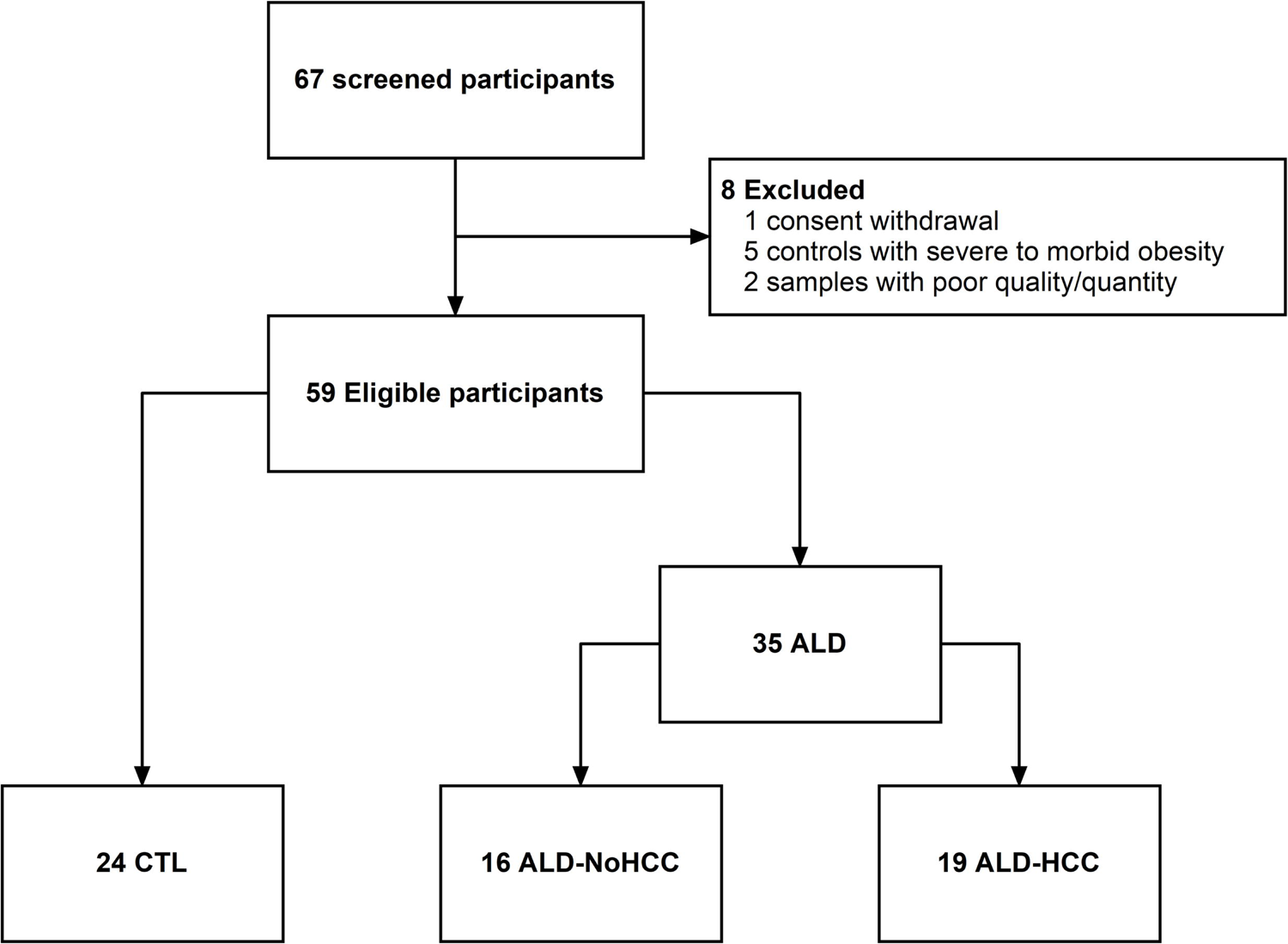
Flowchart of the subjects included in the study. ALD: Alcoholic liver disease; CTL: Controls; HCC: Hepatocellular carcinoma.

**Table 1.** Demographic and clinical characteristics of the study participants.

| Characteristics | Controls<br>(n=24) | ALD-NoHCC<br>(n=16) | ALD-HCC<br>(n=19) | P1 value | P2 value | P3<br>value | P4 value |
| --- | --- | --- | --- | --- | --- | --- | --- |
| Demographic |  |  |  |  |  |  |  |
| Age (years) | 63.82 ± 7.65 | 60.13 ± 10.14 | 65 ± 5.80 | 0.146<sup>M</sup> | 0.857<sup>t</sup> | 0.110<sup>M</sup> | 0.131<sup>A</sup> |
| Gender (M/F) | 10/14 | 11/5 | 19/0 | 0.093<sup>C</sup> | <0.0001<sup>C****</sup> | 0.0085<sup>C**</sup> | 0.0003<sup>C***</sup> |
| Female sex (%) | 14 (58.33 %) | 5 (31.25 %) | 0 (0 %) | -- | -- | -- | -- |
| Male sex (%) | 10 (41.66 %) | 11 (68.75 %) | 19 (100 %) | -- | -- | -- | -- |
| BMI (kg/cm2) | 25.2 ± 2.5 | 27.5 ± 5.98 | 26.89 ± 4 | 0.137<sup>t</sup> | 0.151<sup>t</sup> | 0.721<sup>t</sup> | 0.261<sup>A</sup> |
| Co-morbidities |  |  |  |  |  |  |  |
| Hypertension (%) | 4 (17 %) | 6 (37.5 %) | 12 (63.16 %) | 0.159<sup>F</sup> | 0.004<sup>F**</sup> | 0.181<sup>F</sup> | 0.0074<sup>C**</sup> |
| Dyslipidemia (%) | 0 (0%) | 1 (6.25 %) | 3 (15.79 %) | 0.4<sup>F</sup> | 0.079<sup>F</sup> | 0.608<sup>F</sup> | 0.123<sup>C</sup> |
| Smoking (%) | 0 (0%) | 13 (81.25 %) | 6 (31.58 %) | <0.0001<sup>F****</sup> | 0.005<sup>F**</sup> | 0.006<sup>F**</sup> | <0.0001<sup>C****</sup> |
| Digestive symptoms | 4 (17 %) | 4 (25 %) | 4 (21.05 %) | 0.691<sup>F</sup> | >0.9999<sup>F</sup> | >0.9999<sup>F</sup> | 0.811<sup>C</sup> |
| Liver functions |  |  |  |  |  |  |  |
| AST (U/L) | NA | 54.94 ± 28.10 | 58.95 ± 38.23 | -- | -- | 0.980<sup>M</sup> | -- |
| ALT (U/L) | NA | 90.63 ± 223.14 | 34.79 ± 17.01 | -- | -- | 0.838<sup>M</sup> | -- |
| GGT (U/L) | NA | 192.88 ± 305.55 | 267.21 ± 377.53 | -- | -- | 0.282<sup>M</sup> | -- |
| TBIL (umol/L) | NA | 36.51 ± 56.06 | 25.28 ± 15.04 | -- | -- | 0.898<sup>M</sup> | -- |
| ALP (U/L) | NA | 154.13 ± 60.59 | 177 ± 96.64 | -- | -- | 0.574<sup>M</sup> | -- |
| Serum albumin (g/L) | NA | 33.48 ± 6.36 | 34.91 ± 4.96 | -- | -- | 0.482<sup>t</sup> | -- |
| Routine tests |  |  |  |  |  |  |  |
| Prothrombin time (%) | NA | 67.93 ± 22.49 | 74.35 ± 18.13 | -- | -- | 0.379<sup>t</sup> | -- |
| PLT count (g/L) | NA | 149.75 ± 82.22 | 163.72 ± 87.21 | -- | -- | 0.664<sup>M</sup> | -- |
| Serum creatinine (umol/L) | NA | 81.13 ± 28.55 | 68.71 ± 15.00 | -- | -- | 0.208<sup>M</sup> | -- |
| Complications |  |  |  |  |  |  |  |
| Ascites | 0 (0%) | 6 (37.5 %) | 6 (31.58 %) | -- | -- | 0.736<sup>F</sup> | -- |
| Encephalopathy | 0 (0%) | 7 (43.75 %) | 4 (21.05 %) | -- | -- | 0.273<sup>F</sup> | -- |
| Cirrhosis | 0 (0%) | 14 (87.5 %) | 19 (100%) | -- | -- | 0.202<sup>F</sup> | -- |
| CTP-A | 0 (0%) | 8 (50 %) | 11 (57.89 %) | -- | -- | 0.740<sup>F</sup> | -- |
| CTP-B | 0 (0%) | 7 (43.75 %) | 8 (42.11 %) | -- | -- | >0.9999<sup>F</sup> | -- |
| CTP-C | 0 (0%) | 1 (6.25 %) | 0 (0%) | -- | -- | 0.457<sup>F</sup> | -- |

Continuous variables were expressed as means ± standard deviation. BMI: Body mass index; ALT: Alanine aminotransferase; AST: Aspartate aminotransferase; GGT: Gamma-glutamyltransferase; TBIL: Total bilirubin; PLT: Platelets; ALP: Alkaline phosphatase; CTP: Child-Turcotte-Pugh score; ALD: Alcoholic liver disease; HCC: hepatocellular carcinoma; P1: Comparisons between ALD-NoHCC and controls groups; P2: Comparison between ALD-HCC and controls groups; P3: Comparison between ALD-HCC and ALD-NoHCC groups; P4: Comparisons among all groups; <sup>M</sup>: Mann-Whitney U test; <sup>t</sup>: Unpaired *t*-test; <sup>A</sup>: ANOVA statistical test; <sup>C</sup>: Chi-square test and <sup>F</sup>: Fisher's exact test. \*\* *p* < 0.05, \*\*\* *p* < 0.001, \*\*\*\* *p* < 0.0001. NA: not available.

### Four genera associated with ALD by culturomics

Overall, twenty-one samples were analyzed using microbial culturomics (5 ALD-HCC, 6 ALD-NoHCC, and 10 CTL), allowing the isolation and identification of 32,088 colonies (Table S1). For patients with ALD, 17,308 colonies were isolated and tested using MALDI-TOF MS with an average of 1,573 ± 333 colonies per sample. For control samples, 14,780 colonies were isolated, with an average of 1,478 ± 265 colonies per sample (p = 0.48).

Diversity at the species level was found to be increased in the ALD group for four bacterial genera, all belonging to one phylum (*Bacillota*), including *Thomasclavelia* (p = 0.0016), *Enterocloster* (p = 0.0058), *Clostridium* (p = 0.0021) and *Peptoniphilus* (p= 0.0044, Figure 2a).

**Figure 2.**
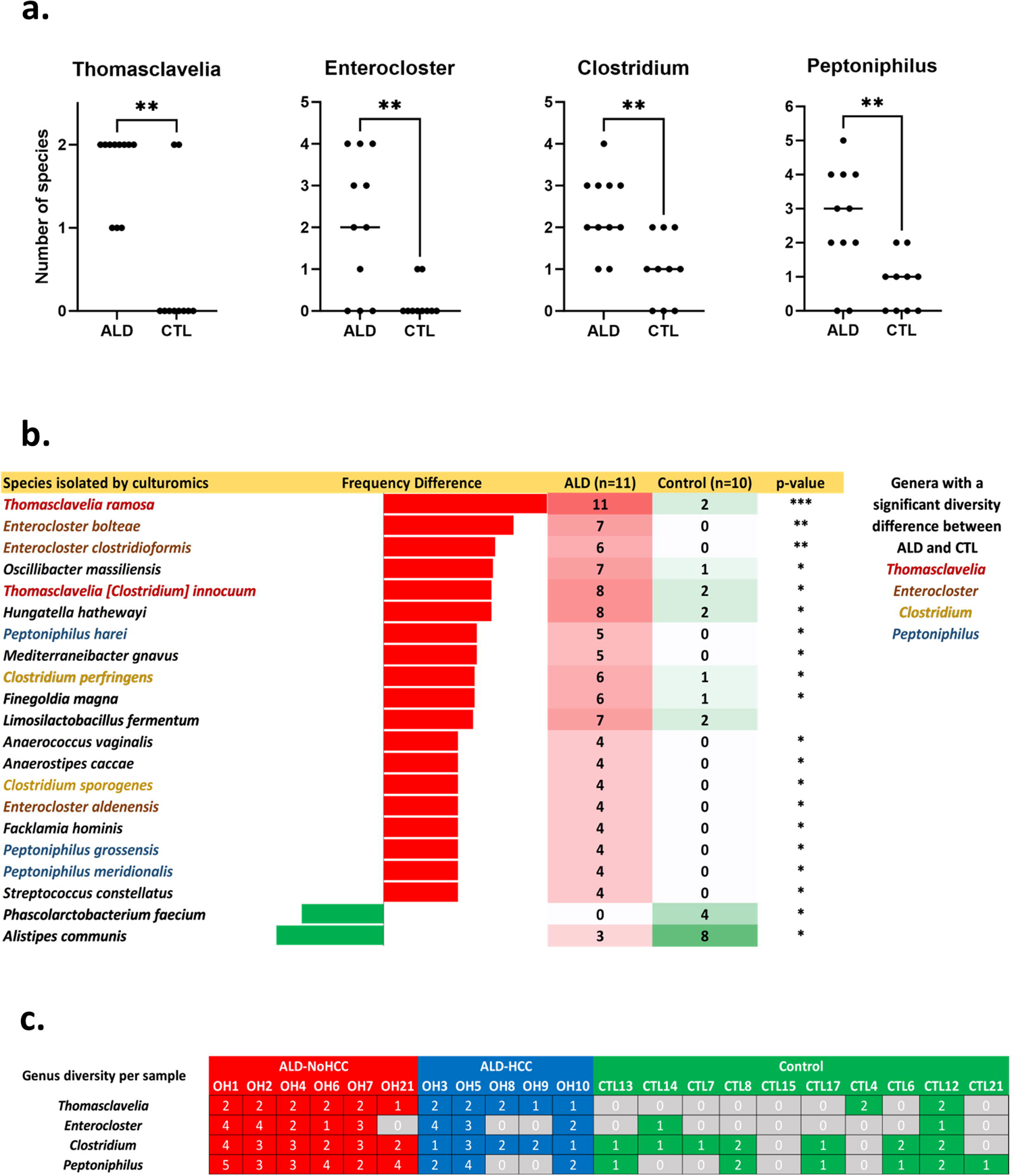
Culturomics results evidenced *Thomasclavelia ramosa* as the gut bacteria the most enriched in ALD. **a.** Species diversity by genus (only four genera with a significant difference are shown). b. Species with a significantly different frequency of detection between ALD and controls. Barnard’s bilateral exact test *p < 0.05, **p< 0.005, ***p<0.0005. At least two species were enriched in ALD for each of the four genera identified in Figure 2a (*Thomasclavelia, Enterocloster, Clostridium, Peptoniphilus*). c. Number of species for each of the four genera associated with ALD for each participant.

### Thomasclavelia ramosa is the most enriched species in ALD by culturomics

At the cultured species level, surprisingly, *T. ramosa* was detected in all ALD patients (11/11 (100%), including those with ALD-HCC (n= 5) and ALD-NoHCC (n= 6), but in only 20% of the controls (2/10, two-tailed Fisher exact test p = 0.00044, Figure 2b). Moreover, among all the cultured species, *T. Ramos* also had the most significant difference in frequency between cases and controls (100% vs. 20%: +80%). This was consistent with the increased diversity of the *Thomasclavelia* genus, as reported above (Figure 2a).

### Enrichment of Enterocloster species in ALD by culturomics

We recently reported an enrichment in *Enterocloster bolteae* in liver diseases associated with metabolic-associated steatohepatitis (34) and hepatitis B virus (36). Here, we found again an enrichment in *Enterocloster bolteae* in liver disease associated with alcoholism (Figure 2b). This was consistent with the increased diversity of this genus in ALD (Figure 2a), particularly the significant increase of 3 species of this genus, including also *E. clostridioformis* and *E. aldenensis* (Figure 2b & Figure 2c). Strikingly, this association was found only by culturomics but not by 16S rRNA sequencing, as observed in our previous studies (34, 36).

### Overview of gut microbiota shift in ALD by 16S rDNA sequencing

The 16S rDNA gene sequencing was performed on the stool samples from all the participants. The sequencing run expressed good-quality monitoring parameters, yielding a total of 4,329,380 reads (1,246,349 for ALD-NoHCC, 1,544,568 for ALD-HCC, and 1,538,463 for the control group) after excluding low-quality reads. On average, each sample had 73,379 ± 54,786 reads. 2,840,773 clean tags were obtained, of which 1,980 OTUs were matched. Of these OTUs, 1,565 (79 %) were successfully assigned at the phylum level, 886 (45 %) at the genus level, and 493 (25 %) at the species level (Table S2). The overall intestinal microbiota structure significantly differed among the ALD-NoHCC, ALD-HCC, and control groups. The Chao1 index (p = 0.037), Simpson index (p = 0.017), and Shannon index (p = 0.0005), which reflect the alpha diversity, were significantly lower in the ALD-NoHCC and ALD-HCC groups compared to the control group. However, no significant difference was noticed in alpha diversity indices between ALD-NoHCC and ALD-HCC groups. PCoA was performed using Bray–Curtis distance, applying pairwise comparison. The ALD-NoHCC and ALD-HCC groups showed a significantly different pattern compared to the control group (PERMANOVA, p = 0.001; F-value= 2.9824; R^2^=0.0963).

### T. ramosa and Mediterraneibacter gnavus enriched in ALD both by culturomics and 16S rRNA amplicon sequencing

Linear discriminant analysis (LDA) effect size modeling was applied to identify specific bacterial taxa associated with ALD-NoHCC, ALD-HCC, and CTL groups (Figure 3). Compared to the control group, the gut microbiota of all ALD patients showed a significantly increased abundance of *Streptococcus_salivarius/Atribacter_sp223* (OTU39314)*, Escherichia_albertii / Escherichia_coli* (OTU2689), Mediterraneibacter_gnavus (formerly, *Ruminococcus_gnavus* (42)), and *Thomasclavelia_ramosa* (formerly *Clostridium ramosum* (43)). Accordingly, the only two species enriched in ALD by culturomics and 16S rRNA sequencing were *T. ramosa* and *Mediterraneibacter gnavus*.

**Figure 3.**
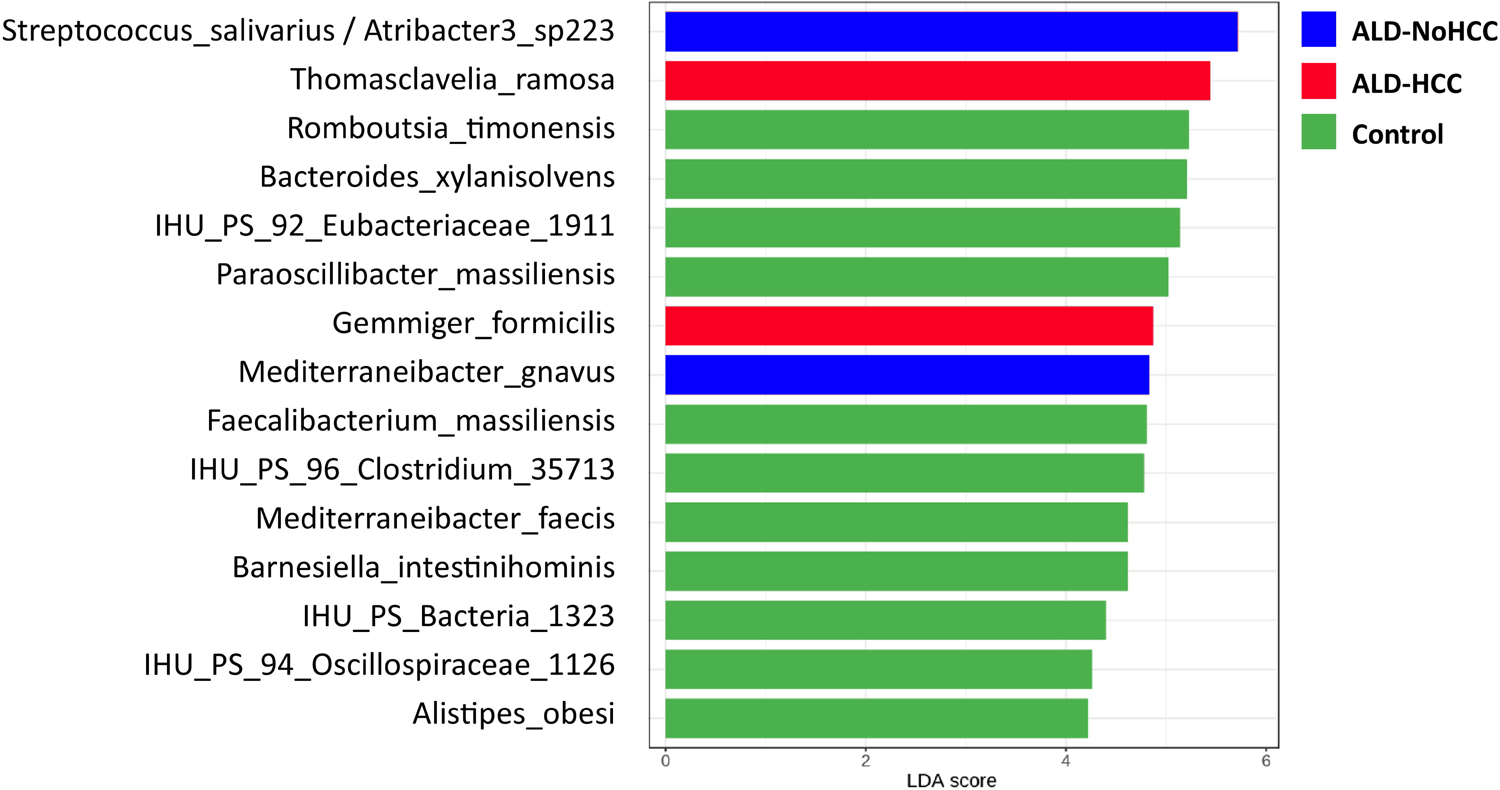
Linear discriminant analysis (LDA) on 16S rRNA sequencing results identified association between *Thomasclavelia ramosa* and liver cancer in alcohol liver disease patients ALD: Alcoholic liver disease; CTL: Controls; HCC: Hepatocellular carcinoma. A logarithmic LDA score > 2 indicated a higher relative abundance in the corresponding group than in other groups.

### Thomasclavelia ramosa was the only species identified by culturomics and sequencing and associated with hepatocellular carcinoma

Including all three groups in a Linear Discriminant Analysis with three groups, the pathobiont *Mediterraneibacter_gnavus* (44, 45) was significantly increased in ALD-NoHCC, while *T_ramosa* and *Gemmiger_formicilis* were the only 2 OTUs significantly associated with the ALD-HCC group (Fig. 3). Accordingly, *T. ramosa* was the only species identified by culturomics and sequencing and associated with hepatocellular carcinoma, identifying it as one of the best oncobiont candidates for further exploration of an instrumental role of a gut microbe for liver cancer associated with alcoholism.

## DISCUSSION

Our study is the first to characterize gut microbiota alteration in ALD and associated HCC using a combined approach of microbial culturomics and 16S rRNA sequencing. The main finding is the identification of *T. ramosa* as an ALD-associated HCC bacterial species.

This signature is supported by the fact that the cases and controls have been certified very strictly with a comparison of groups with homogeneous numbers (around 20 individuals per group), using two different and complementary approaches based on sequencing and microbial culturomics. The robustness of the results obtained by culturomics is based on the number of colonies identified (∼32,000 colonies) and the taxonomic resolution of MALDI-TOF. Identifying ∼1,500 colonies per sample enables robust detection and comparison when a species is not detected (low risk of false negatives). Moreover, the effect size of the association between *T. ramosa* and cancer by 16S rRNA sequencing was high (LDA-score > 5) and very significant.

It has long been known that the intestinal microbiota influences the process of hepatic carcinogenesis via the predominance of certain bacteria (46). Our study identified *Thomasclavelia ramosa* as an oncobiont candidate associated with both ALD and ALD-HCC. The recently revised genome-based bacteria taxonomy classified *T. ramosa* (new genus *Thomasclavelia*) into the family *Coprobacillaceae* within the order *Erysipelotrichales* (43). This is a Gram-positive, non-mobile, spore-forming anaerobic bacillus known for a long time as *Clostridium ramosum* and *Erysipelatoclostridium ramosum* (43). *T. ramosa* is a human pathogen associated with several cases of severe infection (bacteremia, infection of aortic aneurysm, osteomyelitis, arthritis, gas gangrene, Fournier’s gangrene, fatal infections) recently associated with human cancer, notably colorectal cancer (47, 48), but also HCC in a 2023 Chinese study (49). A recent study reported that *T. ramosa* exhibited genotoxicity in an experimental model (50). In this study, screening genotoxic bacteria from patients with inflammatory bowel disease, *T. ramosa* was shown to induce DNA damage and to promote colorectal carcinoma (CRC) tumor growth in mice through small molecules (< 3kDa) production.

The *Enterocloster* genus has recently been defined within the *Clostridium* genus. Interestingly, the two species of this genus associated here with ALD (*E. bolteae* and *E. clostridioformis*) are phylogenetically very close and related to *Enterocloster alcoholdehydrogenati* (51), a species isolated very recently from an alcoholic patient and producing a high level of the liver carcinogen acetaldehyde from ethanol (51, 52). We were among the first to associate *E. bolteae* with liver disease (34, 36), and microbial culturomics was critical as this species was not identified by 16S gene sequencing. Recent trans-omics work has associated this species with liver dysfunction parameters (GGT and ALT (53)). Even if no significant association with cancer has been found in the present study, these species may have contributed to ALD-HCC development in ALD patients.

Limitations of our study included a small sample size. However, the sample size determines the statistical power to identify a statistical significance. However, the clear *T. ramosa* signature identified here means that the difference (effect size) is enormous, that the strength of the association is essential, and that the strength of the association is the first of Bradford Hill’s criteria for causality. Consistency and reproducibility are fulfilled by two previous studies confirming the association with CRC (47, 48) and the confirmation of the *T. ramosa*-liver cancer association by another team (49). Temporality may be confirmed in the future with prospective cohorts of ALD. The linear discriminant analysis fulfills the biological gradient (the higher *the T. ramosa* 16S rRNA number of reads - the higher the risk of HCC). The plausibility and experimental evidence are supported by the reported *in vitro* genotoxicity of *T. ramosa* (50).

The present findings have important clinical implications. At present, besides specific treatment of underlying disease, such as treatment of viral hepatitis or suppression of alcohol consumption, no available treatment prevents HCC development in cirrhotic patients.

Targeting the gut microbiota is a promising approach (54). In the present study, we reported for the first time the association of *T. ramosa* with ALD and, more specifically, ALD-HCC using a combined culturomics and 16S amplicon sequencing approach. Additionally, microbial culturomics allowed us to obtain live bacterial species that will enable the study of mechanisms potentially involved in bacterial-mediated hepatocarcinogenesis (possible liver cell genotoxicity through small molecules production as recently performed by Cao *et al*. (50) for *T. ramosa* and CRC).

Finally, most of the included ALD patients were not drinking anymore at the time of sampling, suggesting the persistence of the ALD signature even after weaning. The persistence of *T. ramosa* may be associated with persistent liver disease progression and cancer risk. In this context, correction of gut dysbiosis represents a new option to reverse liver disease progression and cancer risk after alcohol weaning. Future microbiota-targeted therapeutic perspectives, notably focusing on *T. ramosa,* could reverse the gut dysbiosis-liver vicious circle, thus representing an unexpected hope for preventing HCC in ALD patients after weaning.

## Abbreviations

ALD: Alcoholic liver disease

HCC: Hepatocellular carcinoma

MASH: Metabolic dysfunction-associated steatohepatitis

NGS: next-generation sequencing.

## Authors’ contributions

Conceptualization, funding acquisition, and Project administration: MM, RG. Methodology and visualization: RMW, MM. Resources: AG, PB. Investigation: RMW, AA, BM, AC, CA, GM. Data curation and formal analysis: RMW, AG, AL. Supervision: MTA, MM, RG. Validation: RWM, MM, RG. Writing - original draft: RMW – review & editing: AG, MTA, MM, RG. Reviewed the results and approved the final version of the manuscript: all Authors.

## Data Availability

All data produced in the present work are contained in the manuscript

The authors confirm that the data supporting the findings of this study within the article and/or supplementary materials are available upon request. The raw sequencing data of fecal samples are available in the NCBI Sequence Read Archive with accession number PRJEB62828. The genome of six strains of *Thomasclavelia ramosa* (strains CSUR Q9705, Q9779, Q9849, QA0117, QA0118, and QA0666, available on request) have been sequenced, and genome sequencing data are publicly available under the Bioproject NCBI PRJEB76822.

## Acknowledgments

We thank Vincent Bossi for their excellent technical assistance.

